# Plasma protein LEG1 homolog and post-menopausal breast cancer risk in the UK Biobank

**DOI:** 10.1101/2025.10.26.25338822

**Authors:** Elizabeth A. Platz, Ziqiao Wang, Emily L. Norton, Maria-Eleni Syleouni, Marc J. Gunter, Marcela Guevara, Elio Riboli, Yahya Mahamat Saleh, Karl Smith-Byrne, Vernon A. Burk, Sabine Rohrmann, Nilanjan Chatterjee

## Abstract

We previously identified and confirmed a positive association between plasma protein LEG1 homolog and incident post-menopausal breast cancer risk in two prospective cohort studies that measured the protein using the SomaScan platform. To further confirm this finding in a cohort that used a different proteomics platform, Olink, we investigated this association in the UK Biobank. Among 9,537 post-menopausal, White women followed for a median of 11 years and with protein data, 295 breast cancer cases were identified. Adjusting for breast cancer risk factors, per doubling, the hazard ratio was 1.14 (95% confidence interval CI 0.96-1.35). While the direction of association was consistent, the magnitude was not as strong as in our prior two cohorts (HR per doubling: 1.45, 1.24) and not statistically significant. Data from the UK Biobank provide modest support for plasma protein LEG1 homolog as a risk factor for post-menopausal breast cancer. A quantitative targeted assay is needed.

---

We previously identified a positive association between plasma protein LEG1 homolog (Uniprot Q6P5S2, encoded by *LEG1* and also known as *C6orf58*) and incident post-menopausal breast cancer risk in a prospective cohort analysis in the Atherosclerosis Risk in Communities (ARIC) study (per doubling: HR=1.45, 95% CI 1.23-1.70; p=7.47×10^−6^, false discovery rate adjusted p=0.037, median follow-up was 23 years) (1). The study included 4,403 White (71.9%) and Black (28.1%) middle-aged and older US women, among whom 340 cases were ascertained in 86,335 person-years. To identify plasma proteins associated with breast cancer risk, we individually scanned ~5,000 proteins in relation to breast cancer using Cox proportional hazards models adjusted for demographics and breast cancer risk factors and corrected for false discovery. We confirmed this finding in women in a case-cohort study in the European Prospective Investigation into Cancer and Nutrition (EPIC) study (per doubling: HR=1.24, 95% CI 1.14-1.35; p=9.79×10^−7^, median follow-up was 10 years) adjusting for comparable risk factors (1). In both cohorts, proteins were measured by SomaScan.

In the current study, we sought to confirm protein LEG1 homolog’s association with incident post-menopausal breast cancer risk in a third cohort in which a panel of proteins was measured using a different assay platform. Thus, we conducted a prospective cohort analysis of 295 breast cancer cases among 9,537 post-menopausal, White women in the UK Biobank (2) and who had Olink data for plasma protein LEG1 homolog (OID30608 in Explore 3072). The UK Biobank has received approval from the North West Multi-centre Research Ethics committee. No additional IRB review was needed for this use of de-identified data (https://www.ukbiobank.ac.uk/about-us/how-we-work/ethics/). We excluded women with a prior cancer, who were non-White or related based on genetic data, were not post-menopausal, who did not have proteomic data, or who had missing covariate information (**Figure 1**). We also excluded 76 women (N=17 incident breast cancer cases) who were censored or diagnosed with breast cancer <1 year after study entry because the analysis time scale was age, which is available as an integer. Prior publications reporting on proteomics and breast cancer in the UK Biobank (3, 4) used an earlier proteomic data release that did not include this protein. We used Cox proportional hazards regression with age as the time scale and adjusted for body mass index, height, alcohol drinking (current, former, never), smoking status (current, former, never), hormone replacement therapy use, age at menarche, age at menopause, age at first birth, number of children, and top 10 genetic principal components. We estimated the association with post-menopausal breast cancer per doubling and for quartiles of protein LEG1 homolog.

**Figure 1.**
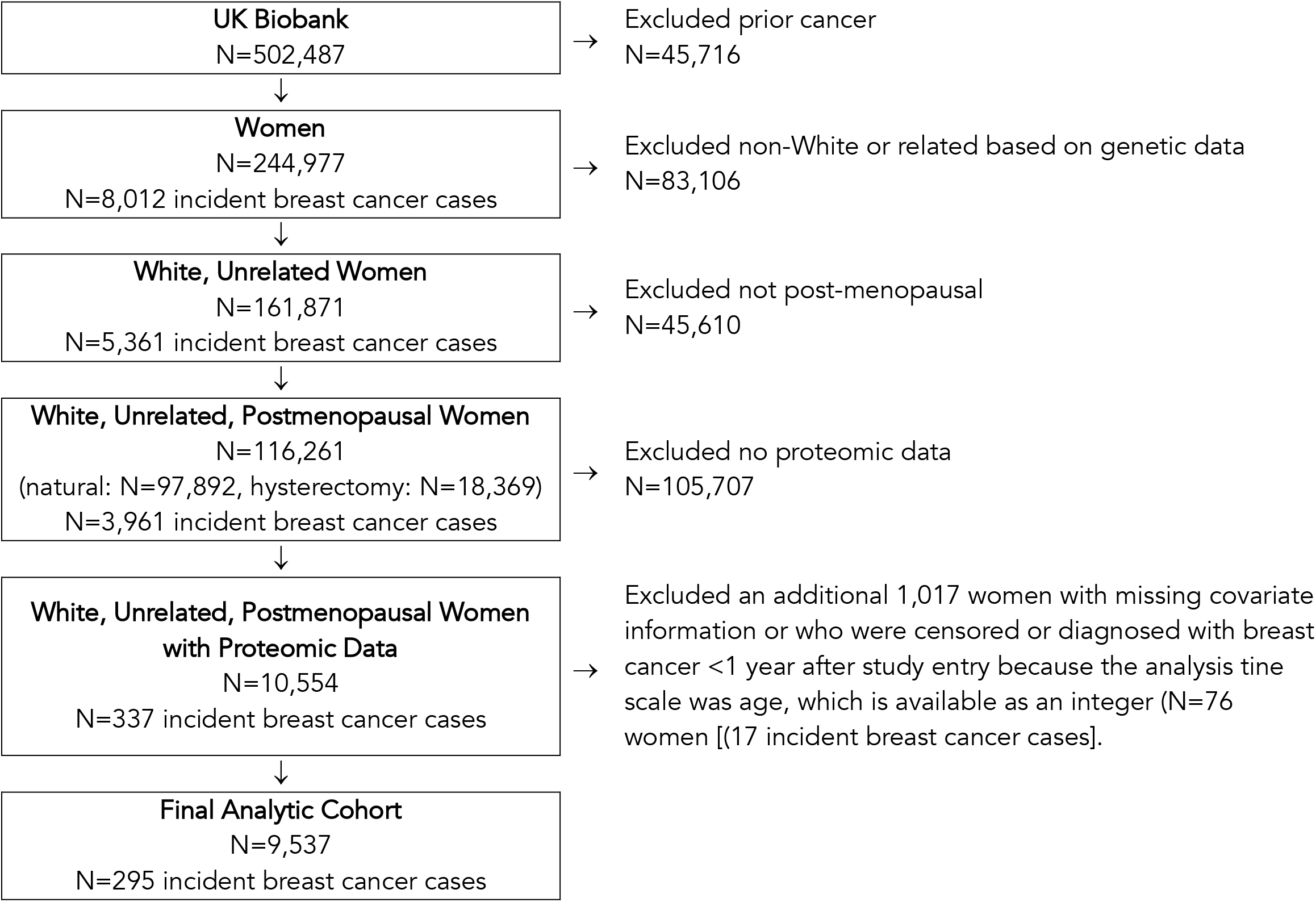
Inclusions and exclusions from the analysis of the association between plasma protein LEG1 homolog and risk of post-menopausal breast cancer, UK Biobank. The final analytic cohort included 9,537 post-menopausal women and 295 incident breast cancer cases. In a sensitivity analysis, we substituted missing age at menopause (for 609 women with natural menopause who did not report age at menopause) and/or missing age at menarche (293 women) with the mode among women in the same quartile for age a blood draw, yielding 10,359 women in the analytic cohort and 318 breast cancer cases.

So that we could compare the association for quartiles among the UK Biobank, ARIC, and EPIC, we re-ran the analysis for EPIC for quartiles of the protein, results which we had not included in our prior publication (1). As before, this analysis included 734 post-menopausal breast cancer cases and subcohort of 2,211 women. The EPIC study was conducted in accordance with the Declaration of Helsinki. The study was approved by the local ethical committees in participating countries and the IARC ethical committee. All participants provided written informed consent for data collection and storage, as well as individual follow-up before study entry. This study is listed at clinicaltrials.gov as NCT03285230.

Per doubling, the multivariable-adjusted HR for the association of protein LEG1 homolog with breast cancer risk was 1.14 (95% CI 0.96-1.35, median follow-up was 11 years). As we observed in ARIC (1), the association was slightly weaker in a minimally-adjusted model (HR=1.11, 95% CI 0.94-1.31, age as the time scale and adjusting for the top 10 genetic principal components). In a sensitivity analysis, we substituted missing age at menopause (609 women with natural menopause who did not report age at menopause) and/or missing age at menarche (293 women) with the mode among women in the same quartile for age a blood draw, increasing the analysis to 318 breast cancer cases in 10,359 women. The multivariable-adjusted HR (1.13, 95% CI 0.96-1.33) was unchanged from overall.

In each of the cohorts – ARIC (from (1)), EPIC, and the UK Biobank – we observed positive direction associations with possible thresholds in quartile analyses (**Table 1**).

**Table 1.** Multivariable-adjusted associations between quartiles of plasma protein LEG1 homolog and post-menopausal breast cancer risk in 3 prospective studies.

|  | Hazard ratio (95% confidence interval) |  |  |
| --- | --- | --- | --- |
|  | UK Biobank*<br>(OID30608) | ARIC**<br>(SeqId_7154_92) | EPIC***<br>(SeqId_7154_92) |
| Quartile 1 | 1.00 (ref) | 1.00 (ref) | 1.00 (ref) |
| Quartile 2 | 1.33 (0.95-1.87) | 1.19 (0.84-1.66) | 1.07 (0.85-1.34) |
| Quartile 3 | 1.33 (0.95-1.86) | 1.76 (1.28-2.41) | 1.36 (1.09-1.70) |
| Quartile 4 | 1.36 (0.97-1.91) | 1.93 (1.40-2.68) | 1.65 (1.31-2.07) |
|  | P=0.12 | P=7.47x10<sup>-6</sup>**** | P=2x10<sup>-6</sup> |

\*Measured using the Olink platform, age as the time scale, and adjusted for body mass index, height, alcohol drinking, smoking status, hormone replacement therapy use, age at menarche, age at menopause, age at first birth, number of children, and top 10 genetic principal components. HRs were nearly identical (1.00, 1.34, 1.33, and 1.37) when omitting the top 10 genetic principal components from the model.
\*\*Previously reported in Syleouni et al. (1), measured using the SomaScan platform
\*\*\*Measured using the SomaScan platform, and adjusted for age at blood collection and recruitment center, height, body mass index, alcohol intake, age at menarche, ever use of hormone replacement therapy at blood collection, diabetes status, first-degree family breast cancer history, ever been pregnant, age at first pregnancy, ever given birth, age at first birth, number of pregnancies, and number of months of breastfeeding.
\*\*\*\*Adjusted for false discovery in the discovery study, P=0.037

While the direction of the association for protein LEG1 homolog and post-menopausal breast cancer in the UK Biobank (HR=1.14) was consistent, the magnitude was not as strong as in ARIC (HR=1.45) or EPIC (HR=1.24) and was not statistically significant. We sought possible explanations for the difference. Each study used a prospective design, restricted to post-menopausal women, had high-quality strategies for case ascertainment and breast cancer risk factor information, and long follow-up. We do not expect that differences in population characteristics (ARIC: US White and Black women; EPIC: women from the United Kingdom, Netherlands, Spain, and Italy; UK Biobank: British women) are explanatory because the associations were comparable in White and Black women in ARIC and roughly comparable between ARIC and EPIC. We also do not expect that our exclusion of 0.8% of otherwise eligible women who had a follow-up time shorter than 1 year in the UK Biobank is explanatory because in ARIC, the 3-year lagged analysis (1.48, 95% CI 1.23-1.77 (1)) the magnitude of the HR was not reduced compared with the main analysis.

A possible explanation is the protein detection technologies. Rooney et al. reported that in a subset of participants in ARIC the Spearman correlation between the measurement of protein LEG1 homolog in Olink and SomaScan was weak (*r*=0.35), and that the coefficient of variation percent for Olink (15.7%) was higher than for SomaScan (7.4%) for LEG1 (5). Thus, it is possible that these proprietary platforms are measuring different features of the same protein (e.g., different epitopes and/or with germline variability in sequence) and/or binding of the detector to the protein with different affinities.

Human protein LEG1 homolog and the gene that encodes it are understudied in general, and in particular, for breast cancer. The *LEG1* gene, also called *C6orf58*, is located on 6q22.33, has 6 exons, and encodes a protein with 330 amino acids (6). The protein appears to be secreted and is evolutionarily conserved (7). With respect to associations for variants in *C6orf58*, the GWAS Catalog indicates 33 reported associations for 25 SNPs in 26 studies with 17 traits, including measures of body weight and height, brain cancer, and traits related to immune activation and elicitors (8). The GWAS Catalog did not include any studies reporting on associations with breast cancer risk. Based on TCGA data, the Human Protein Atlas indicates that the protein is enriched in pancreatic and stomach adenocarcinomas, but not in invasive breast cancer and is not prognostic (9). Thus, at this time no other evidence corroborates a link between this protein and breast cancer.

With respect to plasma levels of protein LEG1 homolog, a study in the UK Biobank reported that variants in *LEG1* (rs62437083-C [intron, minor allele frequency (MAF)<0.05 (6)], rs71565654-C [intergenic, MAF<0.05], rs76226672-G [intergenic, MAF W=0.04, Afr=0.16], rs77864086-A [intergenic, MAF<0.05], rs79111696-C [intergenic, MAF<0.05]) are associated with levels in blood (10), but the minor allele frequencies were low. In ARIC, *cis*-pQTLs were not identified for this protein (11), and for Olink, this protein was not included in a comparable analysis in other cohorts (12). The low minor variant allele frequencies and lack of *cis*-pQTLs precludes analyses of genetically predicted levels in relation to breast cancer risk.

We conclude that data from the UK Biobank provide additional modest support for plasma protein LEG1 homolog as a risk factor for post-menopausal breast cancer. An evaluation of the differences in the detection of this protein by the two platforms is needed, along with the development of a quantitative targeted assay. Mechanistic studies are needed to investigate whether the positive link between protein LEG1 homolog and post-menopausal breast cancer is causal.

## Acknowledgments

*UK Biobank*. This research was conducted using the UK Biobank Resource under Application Number 176948.

*EPIC*. The authors thank all study subjects for their participation and all interviewers who participated in the fieldwork studies in each EPIC center. The authors also thank Bertrand Hemon at IARC for his valuable work and technical support with the EPIC database.

## IARC Disclaimer

Where authors are identified as personnel of the International Agency for Research on Cancer / World Health Organization, the authors alone are responsible for the views expressed in this article and they do not necessarily represent the decisions, policy or views of the International Agency for Research on Cancer / World Health Organization.

## Data Availability Statement

*UK Biobank*. Data from the UK Biobank are available by application to the UK Biobank (https://www.ukbiobank.ac.uk/about-us/how-we-work/access-to-uk-biobank-data/).

*EPIC*. For information on how to submit an application for gaining access to EPIC data and/or biospecimens, please follow the instructions at https://login.research4life.org/tacsgr0epic_iarc_fr/access/index.php

